# Price elasticity of demand for new and recurring opioid prescriptions

**DOI:** 10.1101/2022.01.11.22268987

**Authors:** Rebecca Arden Harris

## Abstract

**Objective:** Several U.S. states have recently enacted excise taxes to curb prescription opioid use and other states are considering similar measures. We assessed the effects of increasing out-of-pocket costs (OPC) on new and recurring opioid fills.

**Methods:** We conducted a retrospective cohort study of opioid-naive individuals presenting with acute back pain using data from a nationwide claims repository. We estimated the effect of OPC on the initiation of opioid treatment in logistic regressions, controlling for socio-demographics, medical history, healthcare utilization, insurance type, and region. With the same covariates plus morphine milligram equivalents and days supplied, we estimated the effect of OPC on the number of opioid fills in negative binomial regressions. We report the price elasticity of demand (PED) for prescription opioids, defined as the percentage change in outcome resulting from a two-fold increase in OPC.

**Results:** Of 25,531 adults diagnosed with acute back pain in Q1 of 2018, 2,451 (9.6%) filled at least one opioid prescription. In multivariable regression, the association between OPC and initiating opioid treatment was not significant (PED= -1.9%; 95% CI: -5.5%, 1.7%). However, by region, the PED was -10.3% (95% CI: -18.1%, -2.4%) in the coastal states and 1.6% (95% CI: -2.5%, 5.7%) in the central-southern states. The PED for the number of prescription fills was -3.7% (95% CI: -7.3%, -0.1%), which also differed by region. In the coastal states, the PED was -15.2% (95% CI: -24.7%, -5.7%) and in the central-southern states -1.5% (95% CI: -5.4%, 2.4%).

**Conclusions:** Opioid fills were price sensitive in the coastal states but not in the central-southern states. Policies that would increase OPC might have a restraining effect on opioid consumption in parts, but not all of the U.S.

## Introduction

In the United States (U.S.), the high rate of prescription opioid consumption remains an urgent public health concern (Holmgren et al., 2020). Recent federal and state level proposals have taken a price-based approach to curb excessive use, including price floors, excise taxes and licensing fees that would raise patients’ out-of-pocket costs (OPC). Price floors can be established directly by insurers or indirectly through governmental policy. The executive branch of the U.S. government has proposed rules to incentivize insurers to raise out-of-pocket opioid prices (Council of Economic Advisers, 2020). State governments can enact opioid excise taxes or licensing fees, as Delaware, Maine, New York, Minnesota, and Rhode Island have done (Harris et al., 2022). Other states are debating similar measures, aiming to raise revenue for treatment and educational programs and discourage opioid consumption through higher OPC (Kwon, 2021). Congress also is considering a bill. Senate Bill 1723 would establish a national opioid sales tax of $0.01 per milligram (117^th^ Congress of the United States, 1^st^ Session, 2021). Yet, the extent to which patients’ initial and recurring prescription opioid use responds to OPC is unknown. To quantify the relationship between consumption and price, economists use the *price elasticity of demand* (PED), a unit-free measure defined as the percentage change in the quantity demanded when there is a one percent change in price, holding everything else constant (Krugman & Wells, 2018).

In general, the demand for a good is said to be inelastic when the absolute value of the PED <1.0. That is, changes in price have a relatively small effect on the quantity of the good demanded. But a PED ≥1.0 is seldom found for pharmaceuticals (Cox, 2009); the inelasticity cut point may be closer to 0.30 for prescription drugs (Mendoza, 2020) and 0.33 for illicit drugs (Gallet, 2014). To date, there have been only two studies, Soni (2019) and Einav et al. (2018), investigating the PED for prescription opioids. A third study, Gatwood et al. (2014), combined opioids and NSAIDs into a single class, making the results difficult to interpret. Both Soni and Einav et al. estimated the PED among Medicare patients, but their findings were far apart. Soni reported a PED of -0.89, which is highly elastic by the standard of most prescription medications. Einav et al. found a PED of -0.04, which is near perfect inelasticity. Neither study disaggregated analysis to the regional or state level despite considerable variation in opioid prescribing across the United States (Centers for Disease Control, 2018; Schieber et al., 2019).

Policy makers assume that patients are informed consumers for whom the demand for prescription opioids will fall as OPC increase. Even though opioids are addictive goods, new users may be price sensitive because physiological dependence takes time to develop, i.e., opioid consumption decisions for new users will not depend on past choices (Becker et al., 2017; Hai & Heckman, 2019). The degree to which new users consider an opioid refill non-discretionary (demand insensitive to price) will likely depend on patients’ unfolding experience of pain severity and tolerability, their understanding of the risks involved in continuing opioid therapy, the availability and convenience of effective substitutes, and their income (Wang, 2007).

The present study estimated the PED for prescription opioids among individuals with private health insurance, the most common form of health care coverage in the U.S. (Keisler-Starkey & Bunch, 2020). Our study cohort was composed of adults who were expected to be responsive to opioid prescription costs: individuals with no history of opioid fills who presented to outpatient care with an initial complaint of non-cancer related musculoskeletal back pain. By providing the first evidence of the impact of cost sharing on new and recurring prescription opioid fills in this population, along with an analysis by region, this paper contributes to our understanding of the ways in which demand for prescription opioids is determined and the feasibility of price interventions to reduce demand.

## Methods

### Ethics statement

This study analyzed preexisting, anonymized data. The University of Pennsylvania Institutional Review Board deemed it exempt from institutional review board approval.

### Data source

Data were extracted from the Optum Clinformatics® DataMart (OptumInsight, Eden Prairie, MN), a large administrative private payer claims database of more than 17 million annually enrolled patients. The database consists of inpatient and outpatient claims; International Classification of Diseases (ICD) diagnosis and procedure codes; prescription fill information including National Drug Code number, quantity dispensed and number of days supplied; out-of-pocket costs; physician specialty codes; patient demographics; and health plan enrollment status. The enrollee population is geographically diverse covering all 50 states, and demographically similar to the United States population of privately insured people in terms of age, gender and race (Optum, 2018).

### Inclusion/exclusion criteria

We selected adult patients (age 18 to 64 years) who were continuously insured from January 1, 2017 through December 31, 2018 and prescribed an opioid for acute back pain (ICD-10 M54.x) in the first quarter of 2018. All patients were treated in outpatient settings and acute back pain was the chief complaint (first listed medical diagnosis) in the index visit. The cohort was followed through December 31, 2018.

Patients were excluded if, in the 12 months prior to the initial visit, they had filled an opioid prescription or received a diagnosis of back pain, because these patients were less likely to be experiencing acute pain. We also excluded patients who were prescribed very high potency drugs such as fentanyl or hydromorphone, or large quantities of tablets (i.e., >99th percentile), as these were improbable treatments for patients presenting with acute back pain. We also excluded extreme values (>99th percentile) in morphine milligram equivalents (MMEs) per day, number of opioid fills, days of opioid supply, and out-of-pocket costs as many appeared to be coding errors. A sensitivity analysis was performed in which we included the cases with very high values in these variables.

### Variables

We had two outcomes of interest: (1) initial opioid fill, a dichotomous variable (yes or no) indicating whether a pharmacy dispensed a prescription for an opioid medication within 10 days of the index office visit; and (2) number of opioid fills, defined as the total of pharmacy fills from the index visit to the end of the follow-up period among patients who had an initial fill.

Out-of-pocket costs was our primary explanatory variable. To measure the effect of OPC on the initial opioid fill, we used the OPC of the index office visit — the sum of patient’s copayment, deductible, and coinsurance — as a proxy for prescription OPC as the prescription costs were unavailable in patients without any fills. To measure the effect of OPC on the number of opioid fills, we used the median of each patient’s prescription OPC for opioid fills. A prescription OPC could vary from one fill to the next if there was a switch in the prescribed opioid drug, strength, or quantity of tablets.

Our models included variables to address the possibility of adverse selection. As an individual’s health status is known only to him or her prior to selecting an insurance plan (Rothschild & Stiglitz, 1976), patients who expect that they will have higher health care needs, as well as patients who prefer more health care, are more likely to choose plans with lower OPC. Thus, patients’ health status and insurance preferences could be correlated with both OPC and opioid fills, which would bias the estimate of the OPC effect upward. To control for adverse selection, we measured patients’ comorbidities (Charlson comorbidity index) and health care utilization (number of office visits) in 2017, the year in which their insurance plans were selected, and included them as potential confounders in our statistical models. We also controlled for recent patient antidepressant use (an antidepressant prescription filled in the quarter 4 of 2017) as antidepressant medication usage has been associated with the initiation of opioid therapy (Carnide et al., 2020) and may contribute to adverse selection.

We controlled for patient age, education, income, race/ethnicity, and gender. Income is likely to have a positive effect on the consumption of prescription opioids as higher-income individuals may be less likely to face tradeoffs between necessities such as food and shelter and other goods and services whose consumption is sometimes delayed such as doctor visits and prescription drugs. Education, race and ethnicity are included as covariates in keeping with the prominent “deaths of despair” hypothesis, which suggests that whites, especially working-class whites, have been disproportionately vulnerable to excessive opioid use (Case & Deaton, 2017). We controlled for gender because women use more health services than men and may be more likely to fill an opioid prescription (Bertakis et al., 2020).

We also evaluated the impact of the type of health insurance plan (preferred provider organization, health maintenance organization, exclusive provider organization, point of service, indemnity, other) on PED as cost sharing may vary by plan type. For patients with ≥1 opioid fill, we controlled for median MMEs per day and total days of opioid medication supplied which may affect OPC (Aroke et al., 2018) and number of fills (Harris et al., 2019).

### Statistical Analysis

All analyses were run in Stata Version 17.0 (StataCorp LLC, College Station, Texas).

### Descriptive statistics

Patients were grouped by whether they had an opioid fill (0 vs. ≥1). The patient characteristics were described using percentages for categorical variables (group differences analyzed using chi-square) and means and standard deviations for continuous variables (group differences analyzed using t-tests).

### Regression models to estimate the PED for opioid fills

We used logistic regression to analyze the effect of OPC on the initial opioid fill, controlling for socio-demographics, medical history including radiculopathy, health care utilization, and insurance type. To estimate the effect of OPC on number of opioid fills among those who had at least one fill, we used zero-truncated negative binomial regression controlling for the same covariates plus MMEs per day and days supplied. We included an offset variable to adjust for differences in follow-up times. To quantify the relationship between consumption and price, we used the *price elasticity of demand* (PED x 100), defined as the percentage change in the quantity demanded (our outcome variable) resulting from a two-fold increase in OPC, holding everything else constant (Mankiw, 2019). The PED was calculated using Stata’s post-estimation command (*margins, eyex*), which computes d(logf)/d(logx), where f is the prediction function specified by the estimation equation (StataCorp, 2021).

### Regional analysis

Regional differences in opioid prescribing and consumption behavior for musculoskeletal pain have been observed in the United States (Jami et al., 2021; Davison et al., 2020), as have regional differences in non-narcotic treatments for pain, such as physical therapy, non-steroidal anti-inflammatory drugs (NSAIDs), and muscle relaxants (Machlin et al., 2011; Zimbelman et al., 2010). While the underlying reasons for these geographic patterns are unclear, together these studies suggest that preferences for treating pain may broadly track the current geo-cultural alignments in the United States – i.e., the divergence between the “coastal” (northeast and western) states and the central-southern states (Klaus & Romain, 2021). Therefore, in addition to our pooled analysis, we fit separate regressions for the coastal states (*U*.*S. Census Bureau Regions – Northeast and West*: Connecticut, Maine, Massachusetts, New Hampshire, New Jersey, New York, Pennsylvania, Rhode Island, and Vermont, Alaska, Arizona, California, Colorado, Hawaii, Idaho, Montana, Nevada, New Mexico, Oregon, Utah, Washington, and Wyoming) and central-southern states (*U*.*S. Census Bureau Regions – Midwest and South*: Illinois, Indiana, Iowa, Kansas, Michigan, Minnesota, Missouri, Nebraska, North Dakota, Ohio, South Dakota, and Wisconsin, Alabama, Arkansas, Delaware, District of Columbia, Florida, Georgia, Kentucky, Louisiana, Maryland, Mississippi, North Carolina, Oklahoma, South Carolina, Tennessee, Texas, Virginia, and West Virginia) as they might produce substantially different PED estimates. We used the Chow test to determine whether the difference in the OPC slopes between the two regions was statistically significant (Chow, 1960).

### Post-hoc analysis

We followed a pre-specified analysis plan but made allowances for additional comparisons and analyses (Rosenbaum, 2010). In view of the results of the regression analyses, we investigated whether three common non-narcotic pain relief treatments for acute back pain – physical therapy (identified from procedure codes), NSAIDs, and muscle relaxants – were used as substitutes for opioid medications (Musich et al, 2019). We used logistic regression to analyze the impact of region on the use of potential substitutes, controlling for radiculopathy, an indicator of severe pain in many patients (Berry et al., 2019). We labeled the analysis “*post-hoc*” for clarity.

## Results

In quarter 1 of 2018, 436,498 continuously insured adult patients with back pain met our initial selection criteria. After excluding patients who had a diagnosis of back pain (n=367,753) or an opioid fill (n=43,214) in the previous 12 months, our final study population was 25,531 opioid-naive patients who presented with new back pain (mean age=45.2 years, sd=12.3; 45.5% male). Of these, 2,451 (9.6%) filled an opioid prescription during quarter 1 of 2018.

Table 1 presents the demographic and clinical characteristics for the full cohort. The mean OPC for the index office visit was $52.02 (sd=$58.66). The mean OPC to fill an opioid prescription was $5.74 (sd=$6.92). Among individuals who filled at least one opioid prescription, the mean number of fills per patient was 2.70 (sd=2.5).

**Table 1.**
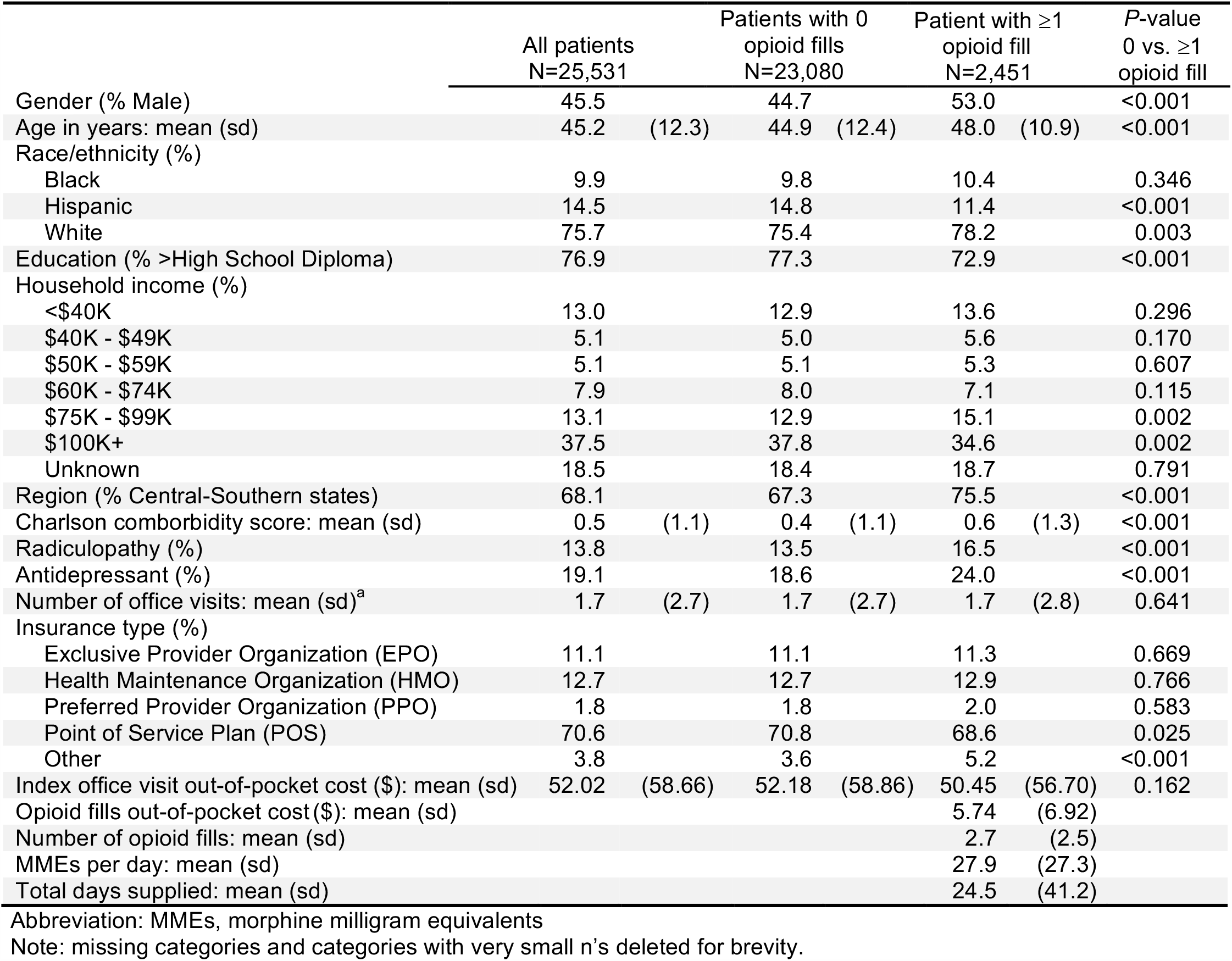
Patient clinical and demographic characteristics associated with an initial opioid fill (0 vs. ≥1)

There were notable differences between the groups of patients based on whether they filled an opioid prescription. Patients without an opioid fill were more often from the coastal states, female, Hispanic or non-White, and slightly younger with more years of formal education and higher income. They also had fewer comorbidities and were less likely to have a diagnosis of acute back pain with radiculopathy or a recent antidepressant prescription fill.

### Impact of OPC on initial opioid prescription fill

Table 2 reports the multivariable logistic regressions. For the full cohort, OPC was not statistically related to initiating opioid treatment (PED=-1.9%; 95% CI: -5.5%, 1.7%; *P*=0.300). Results differed by region, however. The PED was -10.3% (95% CI: -18.1%, -2.4%; *P*=0.010) in the coastal states and 1.6% (95% CI: -2.5%, 5.7%; *P*=0.440) in the central-southern states. The Chow test for the regional interaction effect (OPC x region) was significant (*P*=0.004).

**Table 2.** Regression analysis and price elasticity of demand for an initial prescription opioid fill by U.S. region^a^

|  | All patients<br>N=25,531<br>OR<br>(P-value) | Central Southern<br>states<br>N=17,382<br>OR<br>(P-value) | Coastal<br>states<br>N=8,149<br>OR<br>(P-value) |
| --- | --- | --- | --- |
| **Outcome Variable: Initial opioid fill (0,1)** |  |  |  |
| Gender (Male) | 1.41<br>($<0.001$) | 1.43<br>($<0.001$) | 1.37<br>($<0.001$) |
| Age | 1.21<br>($<0.001$) | 1.19<br>($<0.001$) | 1.27<br>($<0.001$) |
| Race/ethnicity (White) | 1.15<br>(0.005) | 1.12<br>(0.051) | 1.26<br>(0.024) |
| Education (>High School Diploma) | 0.86<br>(0.005) | 0.84<br>(0.002) | 0.97<br>(0.769) |
| Household income: |  |  |  |
| <\$40K | 1.10<br>(0.189) | 1.17<br>(0.064) | 0.90<br>(0.508) |
| \$40K - \$49K | 1.21<br>(0.063) | 1.18<br>(0.164) | 1.33<br>(0.151) |
| \$50K - \$59K | 1.10<br>(0.350) | 1.12<br>(0.347) | 1.07<br>(0.750) |
| \$60K - \$74K | 0.91<br>(0.292) | 0.89<br>(0.264) | 0.98<br>(0.916) |
| \$75K - \$99K | 1.21<br>(0.004) | 1.20<br>(0.017) | 1.24<br>(0.116) |
| Unknown | 1.19<br>(0.007) | 1.19<br>(0.018) | 1.18<br>(0.160) |
| Region (Central-Southern states) | 1.43<br>($<0.001$) | | |
| Charlson comorbidity score | 1.06<br>(0.001) | 1.06<br>(0.004) | 1.07<br>(0.060) |
| Radiculopathy | 1.19<br>(0.004) | 1.24<br>(0.001) | 1.03<br>(0.786) |
| Antidepressant | 1.39<br>($<0.001$) | 1.36<br>($<0.001$) | 1.52<br>($<0.001$) |
| Number of office visits | 0.98<br>(0.055) | 0.99<br>(0.286) | 0.96<br>(0.047) |
| Insurance type: |  |  |  |
| Exclusive Provider Organization (EPO) | 1.04<br>(0.573) | 1.08<br>(0.303) | 0.85<br>(0.346) |
| Health Maintenance Organization (HMO) | 1.00<br>(0.985) | 0.91<br>(0.227) | 1.18<br>(0.179) |
| Preferred Provider Organization (PPO) | 0.91<br>(0.523) | 0.88<br>(0.504) | 0.98<br>(0.946) |
| Other | 0.85<br>(0.070) | 0.99<br>(0.942) | 0.85<br>(0.509) |
| Out-of-pocket cost (\$) for index office visit | 0.99<br>(0.298) | 1.00<br>(0.441) | 0.99<br>(0.010) |
| **Price Elasticity of Demand (PED)** |  |  |  |
| Out-of-pocket office visit costs | -1.9% | 1.6% | -10.3% |
| 95% Confidence Interval | -5.5%, 1.7% | -2.5%, 5.7% | -18.1%, -2.4% |
| P-value | 0.300 | 0.440 | 0.010 |

<sup>a</sup>Multivariable logistic regression models with a binary dependent variable (no opioid fill=0, at least one fill=1). Reference categories for categorical variables: gender, female; race/ethnicity, non-White; education, ≤ high school diploma; income, ≥ \$100K; region, coastal states; radiculopathy, no radiculopathy diagnosis; antidepressant, no antidepressant prescription fill; insurance type, point of service plan (POS). Rounding: ORs that approached but were slightly less than 1.00 were rounded down to 0.99. Abbreviation: OR, odds ratio

### Impact of region and radiculopathy on use of potential substitutes for opioid therapy

Table 3 shows the regional use of physical therapy, NSAIDs, and muscle relaxants in lieu of opioid medications, controlling for radiculopathy. In this *post-hoc* analysis, physical therapy had a strong positive association with radiculopathy (OR=1.39, *P*<0.001) while NSAIDS and muscle relaxants had a strong negative association with radiculopathy (NSAIDS: OR=0.63, *P*<0.001; muscle relaxants: OR=0.49, *P*<0.001). Compared to patients in the coastal states, patients in the central-southern states were less likely to initiate physical therapy instead of opioid therapy (OR=0.71, *P*<0.001), and more likely to initiate NSAIDs (OR=1.29, *P*<0.001) and muscle relaxants (OR=1.47, *P*<0.001).

**Table 3.** Effects of U.S. region and radiculopathy diagnosis on the use of physical therapy, NSAIDS, and muscle relaxants instead of an initial opioid fill (N=25,531)^a^

|  | Odds Ratio | P-value |
| --- | --- | --- |
| **Outcome variable: Physical therapy & no opioid fills (n = 5,989)** |  |  |
| Region: Central-Southern states | 0.71 | <0.001 |
| Radiculopathy | 1.39 | <0.001 |
| **Outcome variable: NSAID & no opioid fills (n = 4,640)** |  |  |
| Region: Central-Southern states | 1.29 | <0.001 |
| Radiculopathy | 0.63 | <0.001 |
| **Outcome variable: Muscle relaxant & no opioid fills (n = 5,738)** |  |  |
| Region: Central-Southern states | 1.47 | <0.001 |
| Radiculopathy | 0.49 | <0.001 |

<sup>a</sup>Multivariable logistic regression models. Reference category for the outcome variables was all other patients in the cohort. Reference category for region was the coastal states. Abbreviation: OR, odds ratio; NSAID, non-steroidal anti-inflammatory drug

### Impact of OPC on number of opioid fills

Table 4 shows that higher out-of-pocket prescription cost was associated with fewer opioid prescription fills among those with ≥1 fill (PED=-3.7%, 95% CI: -7.3%, -0.1%; *P*=0.046). In the coastal states, the association was moderately strong (PED=-15.2%, 95% CI: -24.7%, -5.7%; *P*=0.002). In the central-southern states, the association was not significant (PED=-0.2%, 95% CI: - 5.4%, 2.4%; *P*=0.444). The Chow test for the regional interaction effect (OPC x region) was significant (*P*=0.002). The sensitivity analysis in which we included individuals with very high values yielded similar results.

**Table 4.** Regression analysis and price elasticity of demand for opioid prescription fills among persons with ≥1 fill by U.S. region^a^

|  | All patients<br>N=2,451<br>IRR<br>(P-value) | Central Southern<br>N=1,851<br>IRR<br>(P-value) | Coastal<br>N=600<br>IRR<br>(P-value) |
| --- | --- | --- | --- |
| **Outcome variable: Number of opioid fills** |  |  |  |
| Gender (Male) | 1.08<br>(0.076) | 1.07<br>(0.202) | 1.14<br>(0.131) |
| Age | 1.02<br>(0.404) | 1.01<br>(0.774) | 1.07<br>(0.097) |
| Race/ethnicity (White) | 1.09<br>(0.127) | 1.04<br>(0.514) | 1.29<br>(0.022) |
| Education (>High School Diploma) | 0.97<br>(0.527) | 0.99<br>(0.873) | 0.86<br>(0.204) |
| Household income |  |  |  |
| <\$40K | 1.10<br>(0.228) | 1.01<br>(0.929) | 1.41<br>(0.027) |
| \$40K - \$49K | 1.06<br>(0.552) | 1.02<br>(0.878) | 1.33<br>(0.110) |
| \$50K - \$59K | 1.00<br>(0.966) | 1.03<br>(0.783) | 0.88<br>(0.580) |
| \$60K - \$74K | 0.89<br>(0.234) | 0.89<br>(0.309) | 0.95<br>(0.792) |
| \$75K - \$99K | 1.02<br>(0.749) | 1.01<br>(0.900) | 1.07<br>(0.637) |
| Unknown | 0.94<br>(0.332) | 0.89<br>(0.135) | 1.12<br>(0.350) |
| Region (Central-Southern states) | 1.04<br>(0.484) |  |  |
| Charlson comorbidity score | 1.00<br>(0.818) | 0.99<br>(0.521) | 1.02<br>(0.480) |
| Radiculopathy | 1.24<br>($<0.001$) | 1.27<br>($<0.001$) | 1.14<br>(0.257) |
| Antidepressant | 1.15<br>(0.007) | 1.17<br>(0.010) | 1.08<br>(0.439) |
| Number of office visits | 1.00<br>(0.843) | 1.00<br>(0.987) | 0.99<br>(0.446) |
| Insurance type |  |  |  |
| Exclusive Provider Organization (EPO) | 0.87<br>(0.058) | 0.84<br>(0.038) | 0.99<br>(0.931) |
| Health Maintenance Organization (HMO) | 0.89<br>(0.097) | 0.96<br>(0.608) | 0.79<br>(0.060) |
| Preferred Provider Organization (PPO) | 0.99<br>(0.964) | 0.99<br>(0.950) | 0.87<br>(0.607) |
| Other | 0.86<br>(0.138) | 0.89<br>(0.338) | 0.74<br>(0.191) |
| MMEs per day | 1.01<br>(0.001) | 1.01<br>(0.001) | 1.01<br>($<0.001$) |
| Days supplied | 1.01<br>($<0.001$) | 1.01<br>($<0.001$) | 1.01<br>($<0.001$) |
| Out-of-pocket cost (\$) for opioid fills | 0.99<br>(0.046) | 0.99<br>(0.444) | 0.97<br>(0.002) |
| **Price Elasticity of Demand (PED)** |  |  |  |
| Out-of-pocket prescription costs | -3.7% | -1.5% | -15.2% |
| 95% Confidence Interval | -7.3%, -0.1% | -5.4%, 2.4% | -24.7%, -5.7% |
| P-value | 0.046 | 0.444 | 0.002 |

<sup>a</sup>Zero truncated negative binomial regression models. The dependent variable is a count of the number of opioid prescriptions filled during the observation period.
Abbreviation: IRR, incidence rate ratio

## Discussion

Over the past decade, the United States has experienced substantial human and economic losses from the widespread misuse of opioid medications (McGreal, 2018). Public health interventions, which in many states include restrictions on prescribing (a supply reduction approach), have led to a gradual decline in the number and strength of opioid prescriptions (U.S. Food and Drug Administration, Center for Drug Evaluation and Research, 2020; Cramer et al., 2021; AMA, 2021). Still, each year millions of patients receive opioid prescriptions for pain that is potentially manageable with alternative therapies (Olfson et al, 2020). Legislatures and governmental agencies are now experimenting with price interventions to further deter excessive use and to generate new revenue (a demand reduction approach) (Kwon, 2021). Our findings show that among privately insured, opioid-naive patients with acute back pain, new and recurring opioid fills are responsive to OPC in the coastal states but not in the central-southern states.

In a *post-hoc* analysis, we found that the regional differences in patient use of physical therapy, NSAIDs and muscle relaxants were consistent with the regional differences in opioid price elasticities. Notably, physical therapy was more often prescribed in the presence of severe pain, and more often utilized by patients in the coastal states, suggesting that physical therapy might be viewed as an effective opioid substitute in the coastal states (Frogner et al., 2018). Although this may, in part, explain the sensitivity to prices in the coastal states, our claims data are not specific enough to provide a test of this hypothesis. Contextual information could help elucidate the factors that affect the uptake of physical therapy (Zheng et al., 2017). For example, is there a scarcity of providers and longer wait times in the central-southern states? Are there regional barriers to access in terms of the time commitments and travel distances required of physical therapy? Do physicians and patients in the coastal states have more confidence that physical therapy can alleviate acute back pain? The contours and causes of the use of alternatives to opioid treatment are important areas for future research.

The OPC and PEDs reported in this study can provide some insight into the level of excise taxation that would be needed to lower demand for opioid fills in our cohort of patients. Of the five states that have enacted an opioid excise tax or fee, Delaware and New York defined their tax rates in terms of MMEs, the opioid property that underlies much of the societal cost of prescription drug misuse (Harris et al., 2022). Delaware has a two-tier tax rate that requires manufacturers to pay an impact fee of $0.01 per MME for opioids reported in the state’s Prescription Monitoring Program and $0.0025 per MME for generic substitutes (Del Code, 2021). New York State also imposes a two-tier tax. On the first sale of every opioid unit, the tax rate is $0.015 per MME if the wholesale acquisition cost is ≥$0.50 per unit and $0.0025 per MME if <$0.50 (New York Department of Health, 2020). Maine, Minnesota and Rhode Island imposed annual licensing and registration fees on opioid manufacturers and distributors without an MME multiplier (ME. Rev. Stat., 2021; Minn. Stat., 2021; R.I. Gen. Laws, 2020).

We can see how much tax would be imposed by Delaware and New York by multiplying their tax rates by the MMEs in each prescription filled by our study cohort. Using Delaware’s higher tier rate of $0.01, the tax per prescription would be $0.60 at the 10^th^ percentile (60 MMEs in the prescription fill), $1.50 at the median (150 MMEs), $2.59 at the mean (259 MMEs), and $5.40 at the 90^th^ percentile (540 MMEs). Using New York’s higher tier rate of $0.015, the tax per prescription would be $0.90, $2.25, $3.89, and $8.10, respectively. The tax amounts from Delaware and New York’s lower tier rate of $0.0025 would be much less: $0.15, $0.38, $0.65, and $1.35. Based on our regional PED estimates, the higher tier tax rates of $0.01 to $0.015 would be too low to alter consumption choices in our price insensitive central-southern cohort. In the coastal states, they would reduce new fills by approximately 5-7% and recurring fills by 7-10%. The lower tier tax rate of $0.0025 would have a negligible effect on fills. Importantly, these projections do not take pass-through adjustments into account. The specific tax burdens placed on consumers may vary for the reason that manufacturers, wholesalers, pharmacy benefit managers, insurers and retailers can elect to absorb, spread to other customers, or pass on all or just a portion of the tax to patients. Much of the price-setting process, which involves discounts, rebates, and concessions to supply chain entities, is undisclosed and poorly understood (Ryan & Sood, 2019).

This study has limitations. First, patients with commercial insurance are not representative of all patients. Replicating this research in other populations, such as those with public insurance or no insurance, is warranted. Similarly, a study population enrolled in health plans provided by a single insurer (UnitedHealthcare) may not generalize to all employer health plans. Second, the prescription claims data are for medications dispensed (pharmacy fills); we have no indication as to whether patients consumed the drugs as directed. Third, we used a proxy measure of out-of-pocket costs in our analysis of new opioid fills because most patients did not have an initial fill. Although the proxy demonstrated predictive validity, its precision remains uncertain. Fourth, we had no measure for over-the-counter pain medications, such as acetaminophen and NSAIDs which are widely used (Peck et al., 2021).

The strengths of the study include a large opioid-naïve study population for whom we identified both new and recurring opioid fills. Additionally, to our knowledge, this is the first study to estimate the price elasticity of demand for prescription opioids in a privately insured, opioid-naive population; to model price effects on the initial and subsequent opioid fills; and to analyze the effect of cost-sharing on opioid consumption by region.

## Conclusion

In this population of opioid-naïve patients presenting with new back pain, initial and recurring opioid fills were price sensitive in the coastal states but not in the central-southern states. These findings highlight the need to understand more fully the durable demand for opioid analgesics as the first-line treatment for acute musculoskeletal pain and the feasibility of price interventions to reduce demand.

## Data Availability

All data produced in the present work are contained in the manuscript

## Acknowledgements

This study was part of a master’s degree thesis completed at the University of Pennsylvania. The author would like to thank her thesis advisors – Robert Gross (thesis supervisor), Henry R. Kranzler, and Kyong-Mi Chang – for their guidance and encouragement. The author also would like to thank David S. Mandell, Hillary R. Bogner, Peter W. Groeneveld, and Mark V. Pauly for their many helpful comments and suggestions on earlier drafts of the manuscript. The Leonard Davis Institute of Health Economics at the University of Pennsylvania provided access to the Optum research data base.

## References

1. 117^th^ Congress of the United States, 1^st^ Session. (2021). Senate Bill 1723: Budgeting for Opioid Addiction Treatment Act. Retrieved 28 July 2021 from https://www.congress.gov/117/bills/s1723/BILLS-117s1723is.pdf

2. American Medical Association (AMA). (2021). 2021 Overdose epidemic report. Retrieved 21 September 2021 from https://end-overdose-epidemic.org/wp-content/uploads/2021/09/AMA-2021-Overdose-Epidemic-Report_92021.pdf

3. Aroke H, Buchanan A, Wen X, Ragosta P, Koziol J, Kogut S. (2018). Estimating the direct costs of outpatient opioid prescriptions: A retrospective analysis of data from the Rhode Island Prescription Drug Monitoring Program. J Manag Care Spec Pharm, 24(3):214–224. doi: 10.18553/jmcp.2018.24.3.214. PMID: 29485950.

4. Becker G, Grossman M, Murphy K. (2017). Rational Addiction and the Effect of Price on Consumption. In Determinants of Health: An Economic Perspective. New York Chichester, West Sussex: Columbia University Press. Retrieved 27 September 2021 from https://mgrossman.ws.gc.cuny.edu/files/2017/06/rationaladdic.pdf

5. Berry JA, Elia C, Saini HS, Miulli DE. (2019). A review of lumbar radiculopathy, diagnosis, and treatment. Cureus, 11(10):e5934. doi: 10.7759/cureus.5934. PMID: 31788391; PMCID: PMC6858271.

6. Bertakis KD, Azari R, Helms LJ, Callahan EJ, Robbins JA. (2000). Gender differences in the utilization of health care services. Journal of Family Practice, 49(2):6.

7. Carnide N, Hogg-Johnson S, Côté P, Koehoorn M, Furlan AD. (2020). Factors associated with early opioid dispensing compared with NSAID and muscle relaxant dispensing after a work-related low back injury. Occupational and environmental medicine, 77(9), 637–647. https://doi.org/10.1136/oemed-2019-106380

8. Case A, Deaton A. (2017). Mortality and morbidity in the 21st century. Brookings Pap Econ Act, 2017:397–476.

9. Centers for Disease Control. (2018). U.S. opioid prescribing rate maps. Retrieved 23 July 2021 from https://www.cdc.gov/drugoverdose/maps/rxrate-maps.html.

10. Chow GC. (1960). Tests of equality between sets of coefficients in two linear regressions. Econometrica, 28:591–605. doi.org/10.2307/1910133.

11. Council of Economic Advisers. (2020). The role of opioid prices in the evolving opioid crisis. 2019. Retrieved 30 October 2020 from https://www.whitehouse.gov/wp-content/uploads/2019/04/The-Role-of-Opioid-Prices-in-the-Evolving-Opioid-Crisis.pdf

12. Cox E. (2009). Why Financial Incentives Aren’t Enough to Move the Needle on Compliance. Am Health Drug Benefits, 2(1):12–3

13. Cramer JD, Gunaseelan V, Hu HM, Bicket MC, Waljee JF, Brenner MJ. (2021). Association of state opioid prescription duration limits with changes in opioid prescribing for Medicare beneficiaries. JAMA Internal Medicine, e214281. https://doi.org/10.1001/jamainternmed.2021.4281

14. Davison MA, Lilly DT, Moreno J, Cheng J, Bagley C, Adogwa O. (2020). Regional variation in nonoperative therapy utilization for symptomatic lumbar stenosis and spondylolisthesis: A 2-year costs analysis. Global Spine J, 10(2):138–147. doi: 10.1177/2192568219844227. PMID: 32206512; PMCID: PMC7076589

15. Del Code Tit. 16 Section 4804B(b)(1)-(2) (2021). Retrieved 8 August 2021 from https://delcode.delaware.gov/title16/c048b/index.html

16. Einav L, Finkelstein A, Polyakova M. (2018). Private provision of social insurance: drug-specific price elasticities and cost sharing in Medicare Part D. Am Econ J Econ Policy, 10(3):122–153. doi:10.1257/pol.20160355

17. Frogner BK, Harwood K, Andrilla CHA, Schwartz M, Pines JM. (2018). Physical therapy as the first point of care to treat low back pain: An instrumental variables approach to estimate impact on opioid prescription, health care utilization, and costs. Health Serv Res., 53(6):4629–4646. doi: 10.1111/1475-6773.12984. PMID: 29790166; PMCID: PMC6232429.

18. Gallet CA. (2014). Can price get the monkey off our back? A meta-analysis of illicit drug demand. Health Econ., 23(1):55–68. doi:10.1002/hec.2902

19. Gatwood J, Gibson TB, Chernew ME, Farr AM, Vogtmann E, Fendrick AM. (2014). Price elasticity and medication use: cost sharing across multiple clinical conditions. J Manag Care Spec Pharm, 20(11):1102–1107. doi:10.18553/jmcp.2014.20.11.1102

20. Hai R, Heckman JJ., A Dynamic Model of Health, Addiction, Education, and Wealth. (2019). Becker Friedman Institute for Research in Economics Working Paper No. 2688384, University of Miami Business School Research Paper No. 2688384. Retrieved 27 September 2021 at SSRN: https://ssrn.com/abstract=2688384 or http://dx.doi.org/10.2139/ssrn.2688384

21. Harris RA, Kranzler HR, Chang KM, Doubeni CA, Gross R. (2019). Long-term use of hydrocodone vs. oxycodone in primary care. Drug Alcohol Depend, 205:107524. doi:10.1016/j.drugalcdep.2019.06.026

22. Harris RA, Mandell DS, Gross R. A National Opioid Tax for Treatment Programs in the US: Funding Opportunity But Problems Ahead. JAMA Health Forum. 2022;3(1):e214316. doi:10.1001/jamahealthforum.2021.4316

23. Holmgren AJ, Botelho A, Brandt AM. (2020). A history of prescription drug monitoring programs in the United States: Political appeal and public health efficacy. American Journal of Public Health, 110(8):1191–1197. doi.org/10.2105/AJPH.2020.305696 PMID: 32552023

24. Jami M, Marrache M, Puvanesarajah V, et al. (2021). Treatment of neck pain with opioids in the primary care setting: Trends and geographic variation. Pain Medicine, 22(3):740–745, https://doi.org/10.1093/pm/pnaa372

25. Keisler-Starkey K, Bunch LN. (2020). Health Insurance Coverage in the United States: 2019. United States Census Bureau Current Population Reports, P60-271. U.S. Government Publishing Office, Washington, DC.

26. Klaus D, Romain W. (2021). The cultural divide. The Economic Journal, 131(637): 2058–2088. doi.org/10.1093/ej/ueaa139

27. Krugman PR, Wells R. (2018). Microeconomics. 5th edition. Worth Publishers, Macmillan Learning. New York.

28. Kwon MM. (2021). Pulling the wrong lever opens a trap door: Using taxes to fight the opioid epidemic. Temple Univ LR, 93(2):343–394.

29. Machlin SR, Chevan J, Yu WW, Zodet MW. (2011). Determinants of utilization and expenditures for episodes of ambulatory physical therapy among adults. Physical Therapy, 91(7):1018–1029.

30. Mankiw NG. (2019). Principles of microeconomics. 9th edition. Cengage Learning.

31. ME. Rev. Stat. Tit. 32, §§ 13724,13800-C (2021). Retrieved 8 August 2021 from http://legislature.maine.gov/statutes/32/title32sec13800-C.html

32. McGreal C. (2018). American overdose: the opioid tragedy in three acts. First edition, 2018.

33. Mendoza RL. (2020). Effects of innovation and insurance coverage on price elasticity of demand for prescription drugs: some empirical lessons in pharmacoeconomics. J Med Econ., 23(9):915–922. doi:10.1080/13696998.2020.1772797

34. Minn. Stat. § 151.065 subdiv. 1. (2021). Retrieved 8 August 2021 from https://www.revisor.mn.gov/statutes/cite/151.065/pdf

35. Musich S, Wang SS, Slindee LB, Keown K, Hawkins K, Yeh CS. (2019). Using pain medication intensity to stratify back pain among older adults. Pain Med., 20(2):252–266. doi: 10.1093/pm/pny007. PMID: 29394401; PMCID: PMC6374135.

36. New York State Department of Health. (2020). Opioid Excise Tax Annual Report Guidance. Revised 7/17/20. Retrieved 1 August 2021 from https://health.ny.gov/professionals/narcotic/docs/opioid_ann_rep_guidance.pdf

37. Olfson M, Wang S, Wall MM, Blanco C. (2020). Trends In opioid prescribing and self-reported pain among US adults. Health Aff (Millwood), 39(1):146–154. https://doi.org/10.1377/hlthaff.2019.00783

38. Optum. (2018). Real world health Care experiences from over 150 million unique individuals since 1993. Retrieved 1 November 2020 from https://www.optum.com/content/dam/optum/resources/productSheets/5302_Data_Assets_Chart_Sheet_ISPOR.pdf.

39. Peck J, Urits I, Peoples S, et al. (2021). A comprehensive review of over the counter treatment for chronic low back pain. Pain Ther., 10:69–80. doi.org/10.1007/s40122-020-00209-w

40. R.I. Gen. Laws § 21-28.10-3 (2021). Retrieved 8 August 2021 from https://casetext.com/statute/general-laws-of-rhode-island/title-21-food-and-drugs/chapter-21-2810-opioid-stewardship-act/section-21-2810-3-determination-of-market-share-and-registration-fee

41. Rosenbaum PR. (2010). Design of observational Studies. Springer.

42. Rothschild M, Stiglitz J. (1976). Equilibrium in competitive insurance markets: an essay on the economics of imperfect information. The Quarterly Journal of Economics, 90(4):21.

43. Ryan MS, Sood N. (2019). Analysis of State-Level Drug Pricing Transparency Laws in the United States. JAMA network open, 2(9), e1912104. doi.org/10.1001/jamanetworkopen.2019.12104

44. Schieber LZ, Guy Jr. GP, Seth P, et al. (2019). Trends and patterns of geographic variation in opioid prescribing practices by state, United States, 2006-2017. JAMA Network Open, 2(3). doi:e190665

45. Soni A. (2019). Health insurance, price changes, and the demand for pain relief drugs: evidence from Medicare Part D. Kelley School of Business Research Paper No 19-4. Retrieved 27 July 2021 from https://papers.ssrn.com/sol3/papers.cfm?abstract_id=3268968

46. StataCorp. (2021). Stata 17 Base Reference Manual. College Station, TX: Stata Press.

47. U.S. Food and Drug Administration, Center for Drug Evaluation and Research. (2020). Report to Congress on Opioid Prescribing Limitations. Retrieved August 1, 2021 from https://www.fda.gov/media/147152/download

48. Wang R. (2007). The optimal consumption and the quitting of harmful addictive goods. The B.E. Journal of Economic Analysis & Policy, 7(1). doi.org/10.2202/1935-1682.1684

49. Zheng P, Kao MC, Karayannis NV, Smuck M. (2017). Stagnant physical therapy referral rates alongside rising opioid prescription rates in patients with low back pain in the United States 1997-2010. Spine, 42(9):670–674. doi: 10.1097/BRS.0000000000001875. PMID: 28441685.

50. Zimbelman JL, Juraschek SP, Zhang X, Lin VW. (2010). Physical therapy workforce in the United States: forecasting nationwide shortages. Physical Medicine and Rehabilitation, 2(11):1021–9. doi: 10.1016/j.pmrj.2010.06.015. PMID: 21093838.

